# Gender Distribution of Organizing Committees and Invited Speakers at the International Conferences on Emergency Medicine

**DOI:** 10.1101/2021.08.15.21262072

**Authors:** Gayle Galletta, Imron Subhan, Priyadarshini Marathe

## Abstract

**Background:** It has been documented that women are under-represented as speakers at emergency medicine conferences globally. This lack of opportunity is likely contributing to the gender gap of women in academic and leadership positions.

**Methods:** The Gender-Specific Issues Special Interest Group (GSI-SIG) of the International Federation of Emergency Medicine (IFEM) has analyzed the gender distribution of invited speakers, plenary speakers, and organizing committees from its last three International Conferences on Emergency Medicine in 2016, 2018, and 2019.

**Results:** Men comprised 75% (range 57-92%) of organizing committees, 69% (67-70%) of plenary speakers, and 78% (range 75-81%) of invited speakers.

**Conclusion:** The percentage of women speakers at IFEM’s International Conferences on Emergency Medicine is low; even below the percentage of women emergency medicine physicians. By understanding these data and their consequences, changes can be made to close this gender gap and create more equitable opportunities for women and their career advancement.

**Strengths and Limitations of this study:**

- The gender breakdown of local organizing committee members and invited speakers at three consecutive International Conferences on Emergency Medicine heavily favoured men over women.
- Having women on the host country’s local organizing committee had no effect on the gender distribution of speakers.
- As these conferences draw speakers and participants from a diverse, international audience, the results are generalizable.
- The organizing committee and speakers’ backgrounds (MD, RN, paramedic) were not delineated, and may have skewed the results.
- Gender non-conforming individuals were not identified.

## Introduction

Recent studies have shown that women are under-represented as speakers at emergency medicine (EM) conferences.^1, 2^ Lack of speaking opportunities can have consequences for promotion and leadership roles and may dissuade women from training in EM due to lack of role models. The International Federation of Emergency Medicine (IFEM) is a federation of over 70 national emergency medicine organizations and has held biennial International Conferences on Emergency Medicine (ICEM) until 2018, when these conferences became annual. The 2020 conference was cancelled due to the global pandemic. The Gender-Specific Issues Special Interest Group (GSI-SIG), an expert group within IFEM that addresses issues related to gender and emergency care, has reviewed the gender breakdown of the last three ICEMs to determine whether gender equity is present with regard to organizing and speaking opportunities.

## Methods

Conference programs from the last three ICEMs (Cape Town, South Africa in 2016, Mexico City, Mexico in 2018, and Seoul, South Korea in 2019) were analyzed to determine the gender distribution of the local organizing committee, plenary sessions, and invited speakers. Session chairs, Special Interest Group (SIG) talks, abstract presentations, pre- and post-conference workshops were not included. On two occasions, the gender of a name could not be determined despite a Google search and assistance of a native speaker. This speaker and organizing committee member were excluded from the analysis. It should be noted that it was not possible to determine if there were gender non-conforming committee members or speakers.

## Results

On average, 75% of organizing committees were men (range 57-92%). This number was skewed by the fact that the 2019 ICEM organizing committee in Seoul, South Korea was comprised of 92% men. There were 26 plenary sessions, of which 69% were given by men (range 67-70%). Of the 805 invited speakers, 78% were men (range 75-81%). (Figure 1). Interestingly, the 2016 ICEM conference in Cape Town included an EM nursing track, of which 13 of the 14 speakers were women.

**Figure 1:**
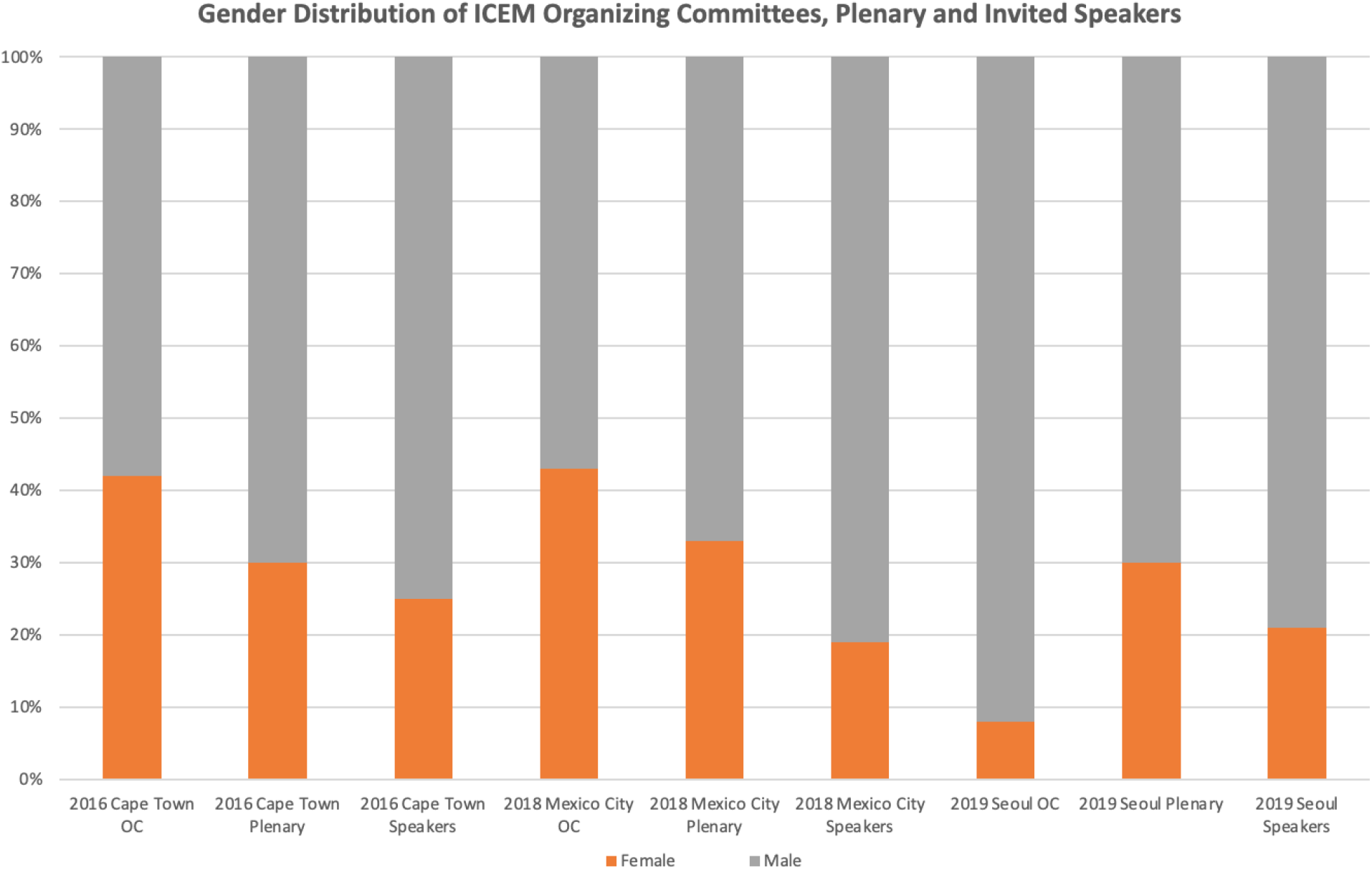
Gender breakdown of Organizing Committees (OC), plenary sessions, and invited speakers at the last three International Conferences on Emergency Medicine.

## Discussion

The percentage of women physicians in general varies greatly among countries, ranging from 20.3% in Japan to 74.3% in Latvia.^3^ The percentage of female EM physicians in particular is thought to range from 28-32% worldwide.^4-6^ In the United States, 38% of current EM residents are female.^4^ Some might argue that the number of female speakers reflects their distribution among the EM specialty. The counterargument is that the number of women at the top, as represented by full professors and Emergency Department (ED) directors, drops off precipitously and could be attributable in part to lack of opportunity and mentorship.^7^ This is often referred to as the “leaky pipe” phenomenon. One previous study found that planning committees with women are more likely to have women as invited speakers.^8^ This was not the case with ICEM. The percentage of women on the organizing committee had little effect on the percentage of women invited as speakers. Furthermore, 13 of the 77 women (16.9%) invited to speak at ICEM Cape Town were in the nursing track. If these female speakers were excluded from the calculations, the results would be skewed even more toward the predominance of male speakers.

Conference organizers should keep gender balance in mind when inviting speakers. This would serve to provide perceptible role models for female physicians and contribute to opportunities needed for career advancement. In addition, organizers should take steps to prevent discrimination against transgender faculty and ensure their fair representation. Specific recommendations to achieve conference speaker gender balance^9^ and the advancement of women physicians in emergency medicine^10^ have already been published.

Within IFEM, there is now an explicit expectation of and strategy to improve gender equity. The ICEM guidelines were updated in April 2019 to require that the local organizing committee “should ensure that there is diversity of speakers, track chairs and conference organizers with no fewer than 40% from either gender. Inclusiveness across gender, race and ableness must be achieved.”^11^ IFEM acts merely in an advisory and oversight role when partnering with the host country’s local organizing committees for the ICEM, and this potentially poses challenges for implementing gender related quotas. Going forward, IFEM’s GSI-SIG will collect and report the data on gender distribution of invited ICEM speakers and is currently developing a speaker policy that can be shared with the organizing committees.

## Conclusions

The vast majority of ICEM organizing committee members, plenary speakers, and invited speakers are men. Some countries and organizations have pledged to have at least 40% of each gender represented in leadership positions.^5^ This percentage has been adopted by IFEM and should be attainable with regards to percentage of invited speakers by gender.

## Data Availability

All data are publicly available in the ICEM program brochures.

## Contributions

GG and PM analyzed the data. IS contributed to the preparation of the document. We would like to thank Dr. Sally McCarthy for her review of the document. The data analyzed are contained in the ICEM program brochures and are publicly available. No patients were involved in this study.

## References

1. Carley S, Carden R, Riley R, et al. Are there too few women presenting at emergency medicine conferences? Emerg Med J. Oct 2016;33(10):681–3. doi:10.1136/emermed-2015-205581

2. Partiali B, Oska S, Touriel RB, Delise A, Barbat A, Folbe A. Gender disparity in speakers at a major academic emergency medicine conference. Emerg Med J. Jan 2020;doi:10.1136/emermed-2019-208865

3. OECD. www.oecd.org. Accessed 05/04/2020,

4. Association of American Medical Colleges. Accessed 4/17/2021,

5. Australian Government Department of Health. Accessed 4/17/2021,

6. General Medical Council. www.gmc-uk.org. Accessed 4/17/2021,

7. Lautenberger D, Dander V, Raezer C, Sloane R. The State of Women in Academic Medicine: the Pipeline and Pathways to Leadership. The Association of American Medical Colleges: www.aamc.org; 2014.

8. Casadevall A, Handelsman J. The presence of female conveners correlates with a higher proportion of female speakers at scientific symposia. mBio. Jan 2014;5(1):e00846–13. doi:10.1128/mBio.00846-13

9. Martin JL. Ten simple rules to achieve conference speaker gender balance. PLoS Comput Biol. Nov 2014;10(11):e1003903. doi:10.1371/journal.pcbi.1003903

10. Choo EK, Kass D, Westergaard M, et al. The Development of Best Practice Recommendations to Support the Hiring, Recruitment, and Advancement of Women Physicians in Emergency Medicine. Acad Emerg Med. 11 2016;23(11):1203–1209. doi:10.1111/acem.13028

11. International Federation of Emergency Medicine. www.IFEM.cc. Accessed 4/20/2021.

